# One Health Intervention Strategies to Address Zoonotic Neglected Tropical Diseases in Southeast Asia: A Scoping Review

**DOI:** 10.1101/2023.09.21.23295764

**Authors:** Agrin Zauyani Putri, Adarsh Varghese George, Shannon A. McMahon, Claire J. Standley

**Affiliations:** Heidelberg Institute of Global Health (HIGH), Heidelberg University, Heidelberg, Germany; John Hopkins Bloomberg School of Public Health, Baltimore, Maryland, United States of America; Center for Global Health Science and Security, Georgetown University, Washington, DC, United States of America

## Abstract

**Background:** Neglected tropical diseases (NTDs) affect over 1 billion people worldwide, causing life-long disabilities and death. Many of the 20 NTDs prioritized by the World Health Organization are zoonotic, spreading between animals and humans. Southeast Asia faces a significant NTD burden, including zoonotic parasitic infections like echinococcosis, taeniasis/cysticercosis, and foodborne trematodiases (FBT). Despite recent research and programs emphasizing the importance of One Health efforts, there is limited insight into their implementation. This scoping review explores existing approaches for multi- and cross-sectoral engagement with respect to three parasitic zoonotic NTDs in Southeast Asia. Additionally, we aimed to synthesize the One Health practices and advocate effective strategies for region-specific collaboration to tackle parasitic zoonotic NTDs.

**Methodology and principal findings:** We utilized the five-step framework of Arskey and O’Malley to conduct this scoping review. We systematically searched Pubmed, Web of Science, and Medline for peer-reviewed literatures. A total of 1725 publications were screened for eligibility and 105 publications identified for full-text screening. Fifteen (15) publications met our inclusion criteria, and we examined them for key themes related to One Health intervention strategies. Five themes emerged from our analysis: 1) health education, 2) treatment, 3) WASH practices, 4) ecosystem monitoring, and 5) surveillance and diagnostics. Additionally, we identified challenges cited during the implementation process, which included limited resources, community engagement, coordination and collaboration, and political commitment.

**Conclusions:** The importance of adopting a comprehensive strategy is strongly supported by the literature and WHO’s emphasis on One Health in the new NTD Road Map. While there are examples of One Health approaches being implemented to control parasitic zoonotic NTDs, the application of One Health strategies is not consistent throughout the Southeast Asia region. Therefore, there are valuable opportunities to develop an implementation research agenda and enhance regional coordination mechanisms. Additionally, future potential strategies should consider determinants of health in NTDs through a multi-sectoral lens.

**Author Summary:** Southeast Asia is among the top-three regions reported requiring interventions against NTDs, including those which can be transmitted between humans and animals (known as zoonotic diseases), such as foodborne trematodiases (FBT), taeniasis/cysticercosis, and echinococcosis. Taeniasis/cysticercosis, a pig-associated zoonotic disease, is a major cause of epilepsy due to brain infection, while FBT is a major cause of aggressive cancer known as cholangiocarcinoma and can also cause pulmonary infections that are usually misdiagnosed. Additionally, echinococcosis is a disease that causes cysts in the liver, lungs, and other organs. Although One Health has been advocated as an important approach to control zoonotic NTDs, there remains limited knowledge regarding the implementation of these approaches in Southeast Asia region. We conducted a scoping review to identify ways in which multisectoral and One Health approaches have been used in Southeast Asia to prevent and control FBT, taeniasis/cysticercosis, and echinococcosis. We found that examples for control of FBT and taeniasis/cysticercosis, but none for echinococcosis, and distribution was not consistent throughout the countries in the region. To this end, we suggest further implementation research as well as leveraging regional coordination mechanisms could be beneficial to further advance One Health as a tool for managing zoonotic NTDs in Southeast Asia.

## Introduction

Neglected Tropical Diseases (NTD) disproportionately affect poor populations and cause chronic life-long disabilities and even death if left untreated (1,2). Despite harming the health, social wellbeing and economic development of more than 1 billion people worldwide (3), these diseases receive comparatively less attention in the global health agenda and little funding and research (4). Among the 20 NTDs that the World Health Organization (WHO) prioritizes, 10 are zoonoses, meaning they can be transmitted between humans and other animals: rabies, scabies, dracunculiasis, trypanosomiasis, Chagas diseases, leishmaniases, leptospirosis, echinococcosis, taeniasis/cysticercosis, and FBT. Zoonotic diseases primarily occur in communities where people work and live closely together with animals (5,6), and can be especially devastating because they not only cause human illness but also harm livestock, thereby reducing a family’s income and sources of nutrition (6). Moreover, in neglected parasitic diseases, the role of animals in transmission is often overlooked even through breaking a parasites’ life cycle is critical to control and eliminate diseases.

To effectively prevent and eliminate NTDs, a range of public health actions, health promotion efforts, and medical interventions are necessary (7,8). Integrated approaches, such as the One Health approach, are needed to address zoonotic diseases through cross-cutting activities that involve multiple sectors (8,9). The One Health approach recognizes the interconnectedness of human, animal, and environmental health, and seeks to address health issues at their intersection and in interdisciplinary manner (10,11). Implementing a One Health approach can help ensure that NTD control and elimination efforts are sustainable and effective (8). In 2020, the WHO developed the NTD Road Map for 2021-2030. This road map seeks to end fragmented implementation of NTD programs by advocating for integrated approaches, such as One Health, through cross-cutting activities involving multiple diseases. The road map provides a guide and resource for countries that are interested in addressing NTDs (12). There are five core strategies that WHO recommends in order to control NTDs: Individual disease management; preventive chemotherapy; vector control; veterinary public health; and water, sanitation, and hygiene (WASH), within a systemic and cross-cutting approach (8).

Southeast Asia is among the top three regions of the world heavily burdened with NTDs (5,13). Of the NTDs present in the region, FBT, taeniasis/cysticercosis and echinococcosis, all neglected parasitic zoonosis, are responsible for a substantial burden of disease in the region (14–17). A severe infection caused by these parasites can be fatal and result in long-lasting problems such as malnutrition, anemia, growth and cognitive impairment, and a decreased ability to tolerate co-infection with other diseases (18–22). Historically, vertical disease control efforts using mass drug administration (MDA) of antihelmintic medication, have been promoted across all endemic regions in order to lower the prevalence of parasitic NTDs infections (23,24). However, these MDA approaches have not mitigated long-term impacts on transmission - despite successfully reducing incidence in the short term. For example, a study in Vietnam showed high reinfection rates within the first year of the last MDA programs for FBT and taeniasis (25). Additionally, reinfections have also been reported in settings that were previously certified as having eradicated guinea worm diseases (GWD)— due to the persistence of animal hosts that maintain transmission (26–29). The complexity of zoonotic parasitic life cycles, which often involve snails as intermediate hosts, animal reservoir hosts, and both animal and human definitive hosts, means that elimination and control of the diseases require a solid One Health approach (30).

There has been a growing number of One Health related global, regional, and national initiatives in Southeast Asia, including some to tackle NTDs in particular. These efforts reflect a growing recognition of the importance of a holistic approach. Notably, four international agencies (WHO, FAO, WOAH, UNEP) have recently developed a collaborative One Health Joint Plan of Action to control and eliminate NZDs. Examples of national initiative can be seen in Indonesia with respect to controlling rabies and foodborne diseases, as well as through the Mekong Basin Diseases Surveillance (MBDS) network as regional cooperation among 6 countries (31).

Despite evidence of the importance of One Health for NTD control, and the existence of projects and policies citing One Health approaches, limited information exists regarding how the strategy can best be delivered to tackle parasitic zoonotic NTDs in Southeast Asia. To address this gap, we conducted a scoping review to explore aspects of multi- or cross-sectoral engagement in controlling echinococcosis, taeniasis/cysticercosis, and FBT in Southeast Asia. We aimed to synthesize effective One Health practices that could be beneficially applied in other countries in the region, and to then highlight effective strategies for region-specific collaboration to tackle parasitic zoonotic NTDs.

## Methods

The scoping review approach was based on the methodological framework by Arksey and O’Malley (32). The five stages of the framework consist of: 1) identifying the research question, 2) identifying relevant studies, 3) selecting studies, 4) charting the data, and 5) summarizing and reporting the findings.

We conducted a scoping review on intervention strategies using the One Heath approach on three zoonotic NTDs known to be of public health importance in Southeast Asia: echinococcosis, taeniasis/cysticercosis, and FBT. Our target study area was Southeast Asia, as defined by membership in the Association of Southeast Asian Nations (ASEAN), namely: Indonesia, Philippines, Malaysia, Vietnam, Cambodia, Thailand, Lao PDR, Myanmar, Singapore, Brunei, and Timor-Leste.

### Identification of relevant studies

The lead author searched PubMed, Web of Science, and Medline using structured search terms: [list of diseases] AND [list country settings] AND [list of integrated/collaborated intervention] (S1 Table). English-language articles published from January 2005 to December 2020 were included if they contained information about interdisciplinary, multidisciplinary, or One Health intervention strategies with respect to the diseases and countries of interest (Table 1). The 2005-2020 window was selected as these dates mark cornerstones for One Health. In 2005, a year after the publication of the “Manhattan Principles on One World – One Health”, the phrase “One Health” entered the scientific and programmatic literature. The final month of 2020 marks a logical endpoint for this research given that, in January 2021, the WHO published their new Road Map on NTDs, which aims to reduce the number of people in need of NTDs treatment, reduce DALYs, and eliminate and eradicate several NTDs by 2030.

**Table 1.**
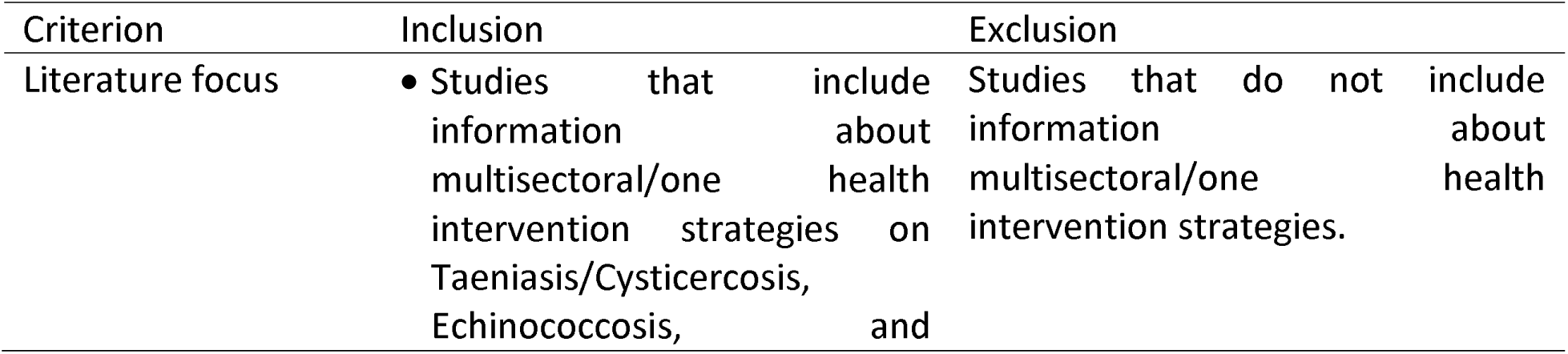
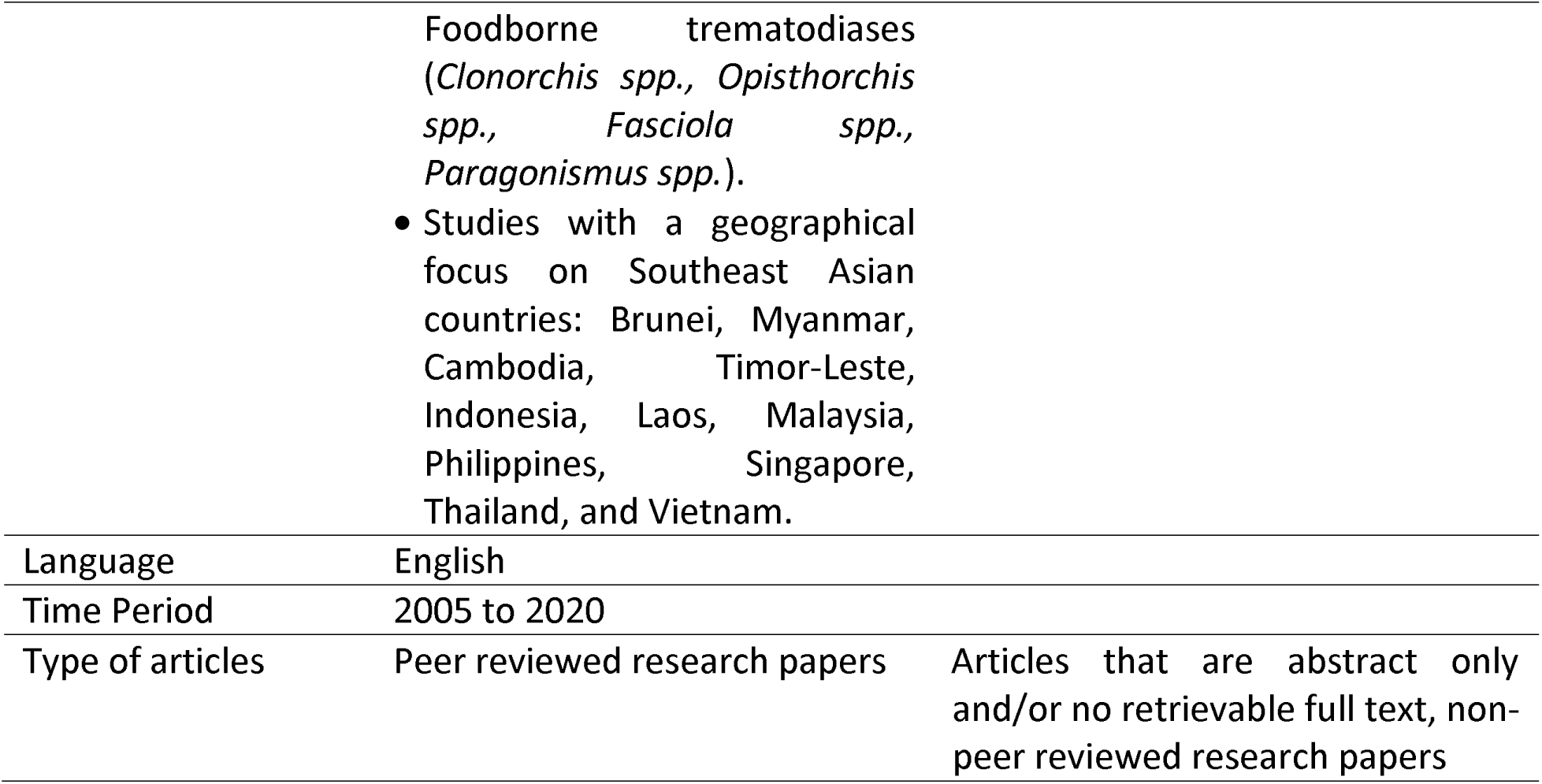
Inclusion and exclusion Criteria.

### Data extraction and data analysis

Two authors screened the papers independently based on the eligibility criteria and conferred on papers to include. All the studies were screened, first on an abstract/title level and the remaining full texts were subsequently read. Finally, the included papers were retrieved for final screening and data extraction. Data were collected and tabulated on a charting table sheet (S2 File) with three types of information related to the: publication, study setting, and intervention strategy. First, information was collected related to a publication’s author, year of publication, title, and objectives of study. Second, details of a study setting were collected. Third, thematic categories relating to intervention strategies, attendant challenges, and study outcomes were derived through the full-text review and data extraction. For each included paper, the findings of the One Health intervention implementations were extracted, with particular attention to how study authors described any results with respect to effectiveness or other outcomes.

## Results

In total 1540 publications were screened and assessed for eligibility based on inclusion and exclusion criteria (Fig 1), of which 105 full-texts were reviewed. A total of 90 full-text publications did not meet the inclusion criteria because their content was outside the scope of interest, abstract only, and/or written in languages other than English, leaving 15 final papers for synthesis.

**Fig 1.**
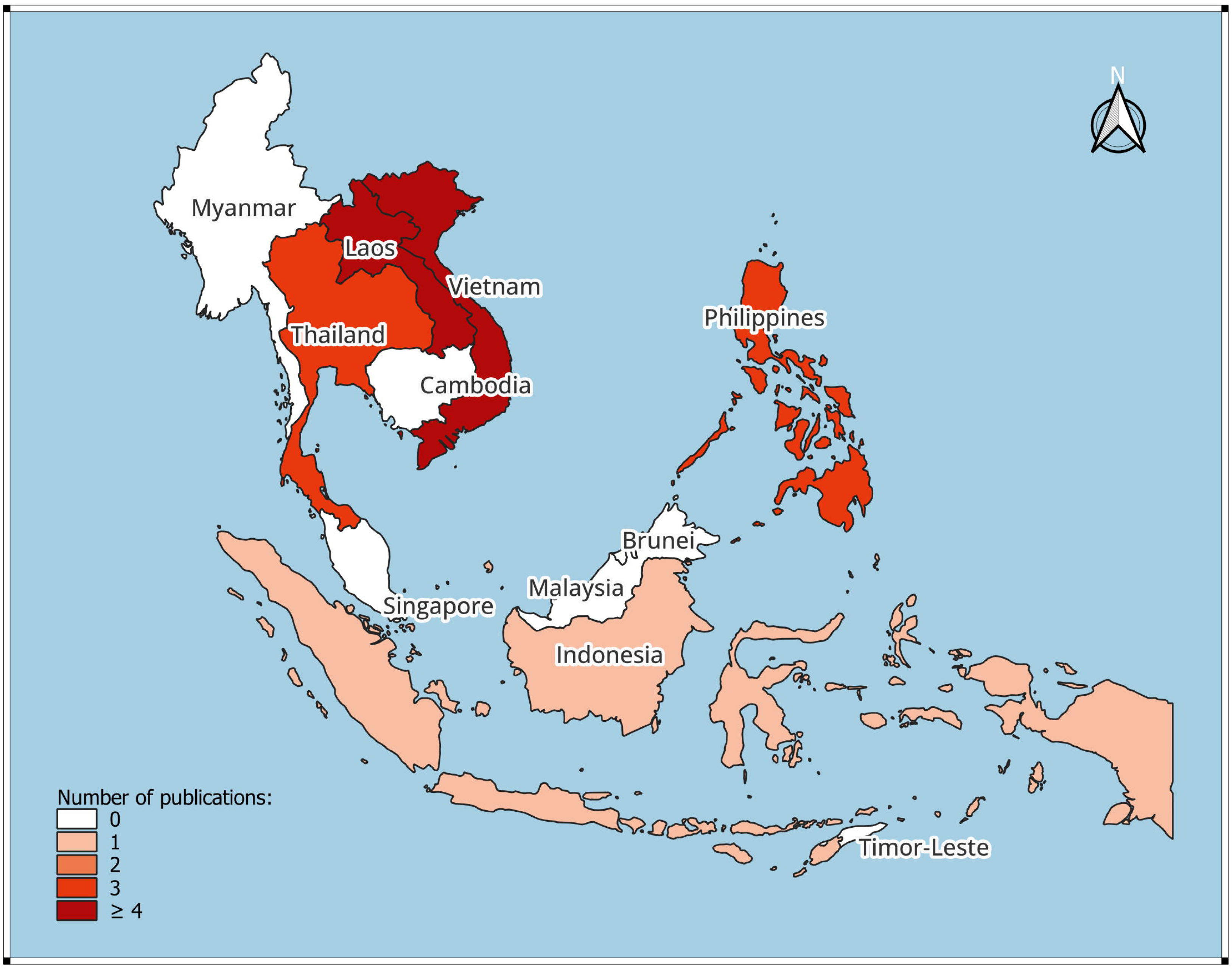
PRISMA diagram of articles identified and selected.

### Characteristics of the included studies

The 15 included studies were conducted in five countries (Fig 2), namely Lao People’s Democratic Republic (Lao PDR) (33–36), Indonesia (37), Philippines (38–40), Vietnam (41–44), and Thailand (36,45,46). Studies focused on control of taeniasis/cysticercosis and FBT (*Clonorchis spp., Opisthorchis spp., Paragonismus spp.*); no study focused on echinococcosis.

**Fig 2.**
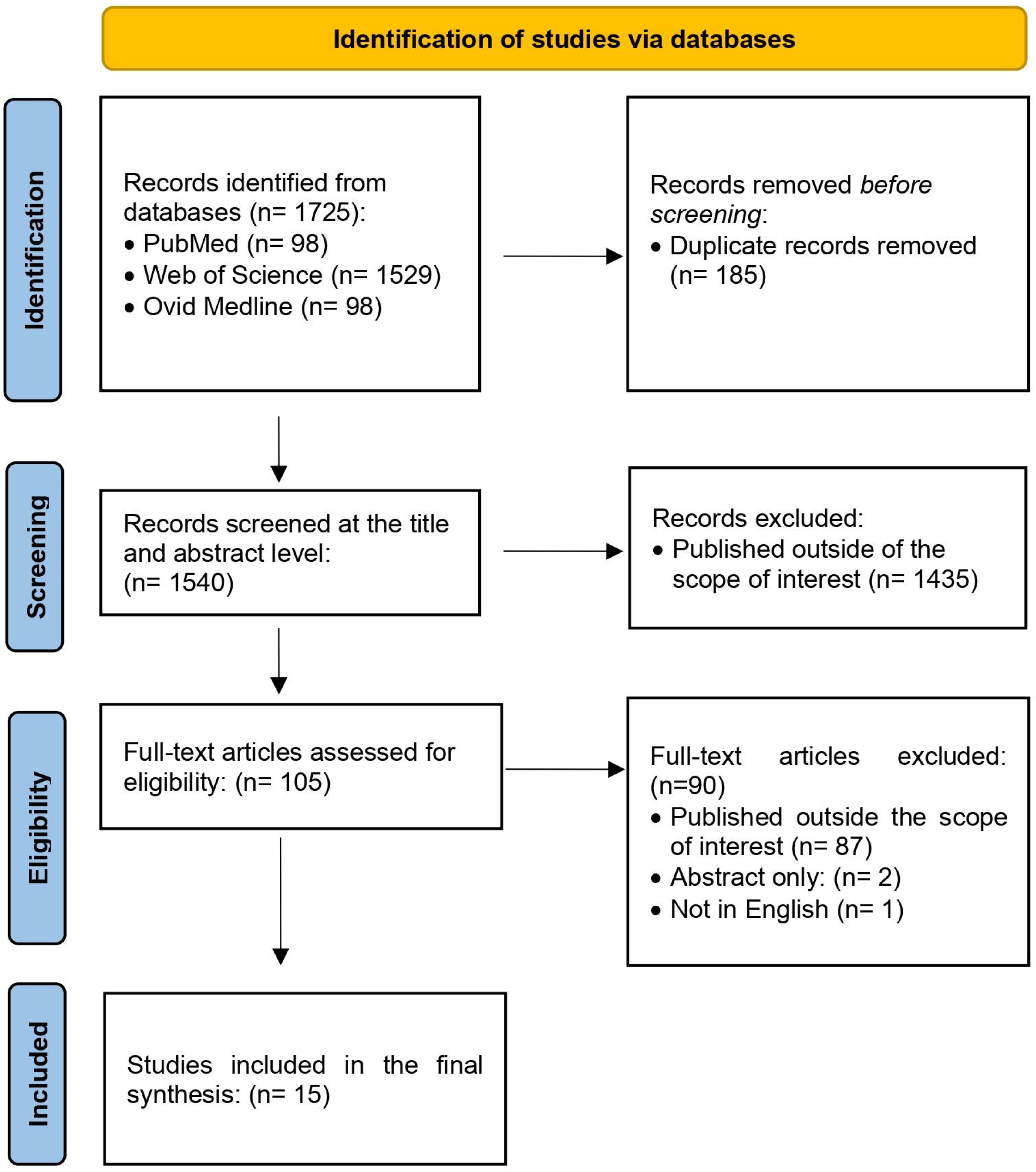
Map of the distribution of included papers in Southeast Asia. Base map layer from Natural Earth (https://naciscdn.org/naturalearth/packages/Natural_Earth_quick_start.zip) and edited with QGIS.

### Key findings

We identified five intervention themes from the included studies that highlight the nature of One Health in the control of taeniasis/cysticercosis and FBT within Southeast Asia. The intervention themes were: health education; treatment; WASH practices; ecosystem monitoring; and surveillance and diagnostics. Each category of the themes was examined based on how the country implements it, multidisciplinary elements, the outcomes for NTD control, and implementation challenges. A summary of the One Health interventions as extracted from the papers is presented in Table 2.

**Table 2.**
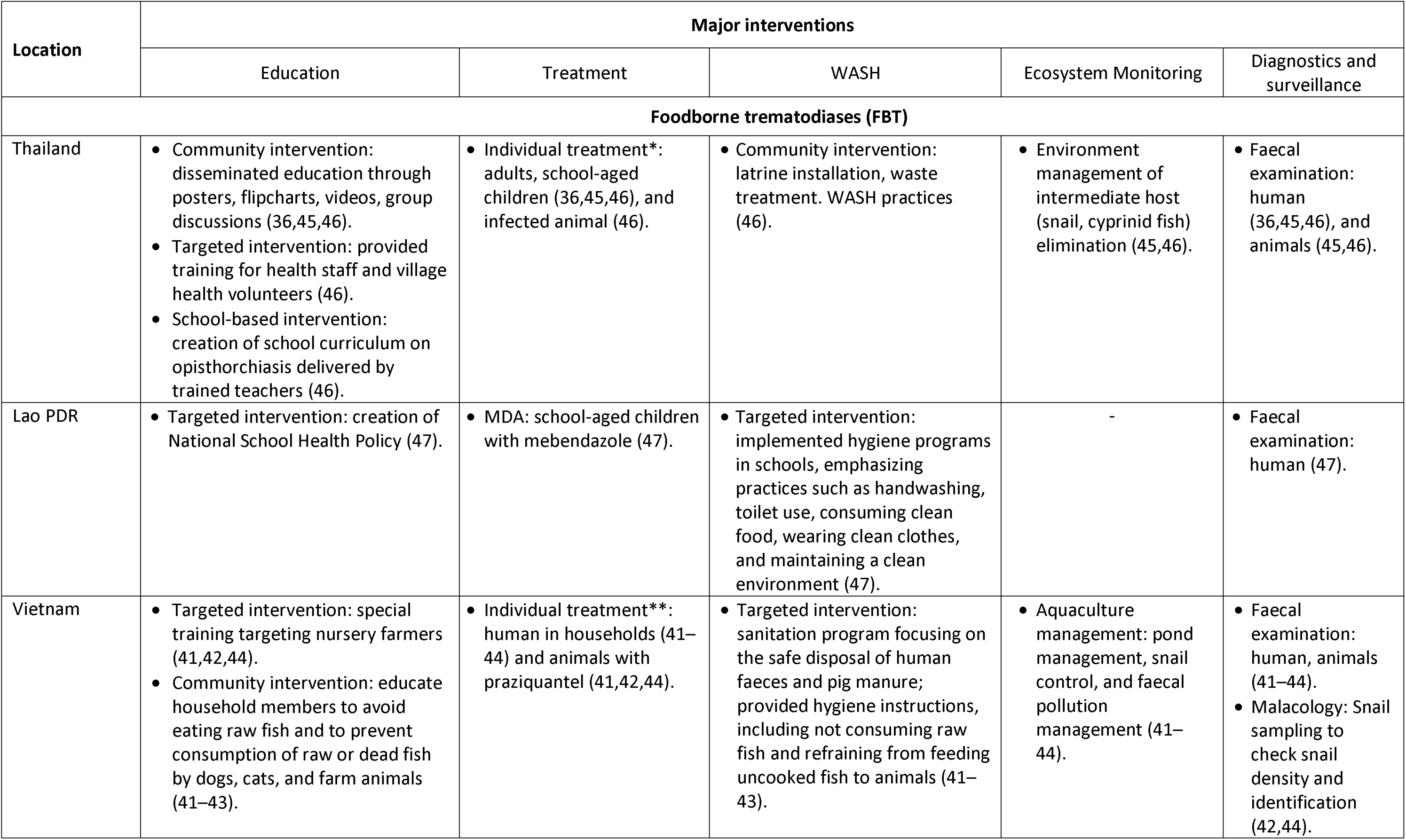
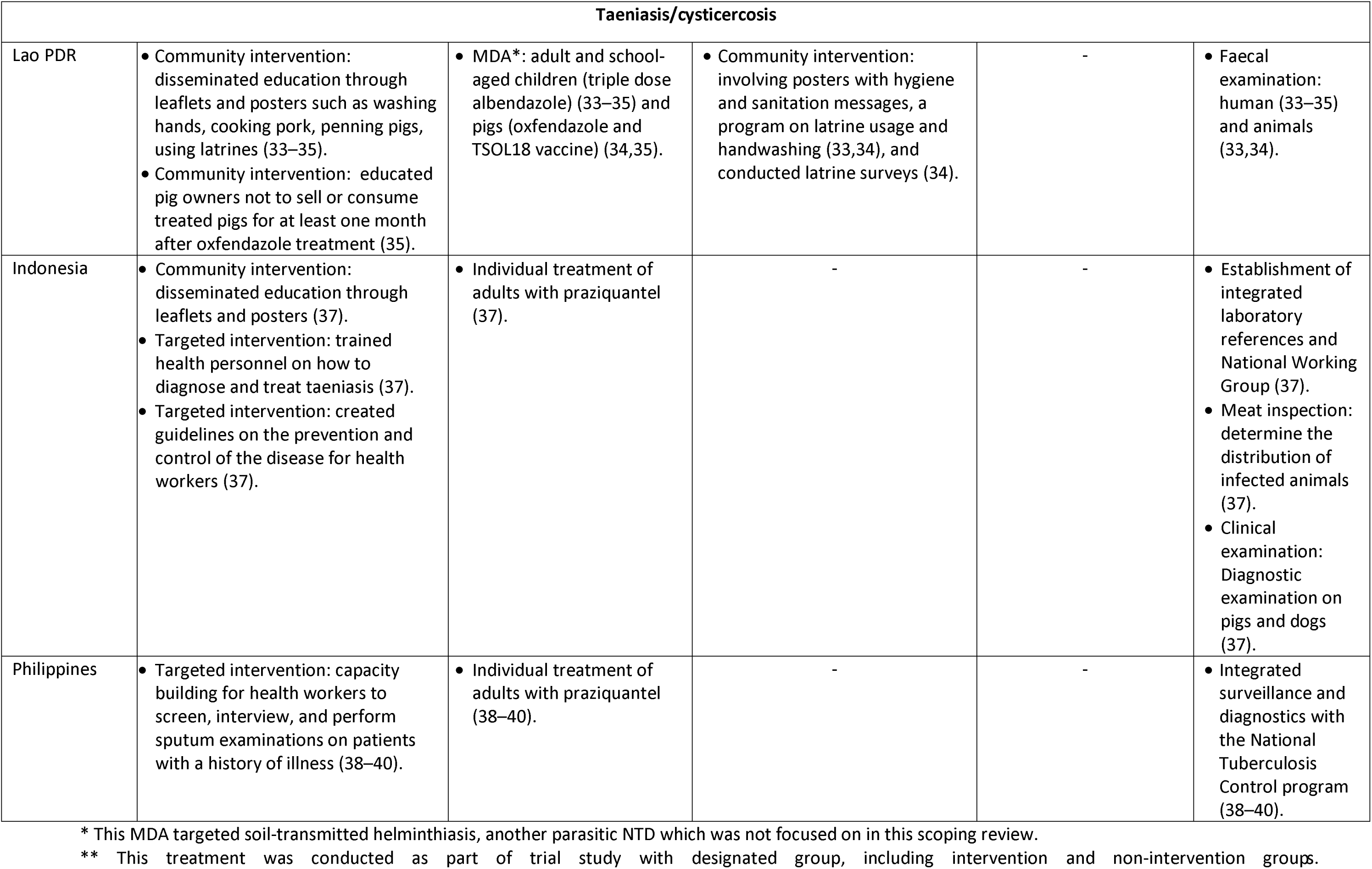
A summary of interventions identified in the included studies.

| Location | Major interventions |  |  |  |  |
| --- | --- | --- | --- | --- | --- |
|  | Education | Treatment | WASH | Ecosystem Monitoring | Diagnostics and surveillance |
| **Foodborne trematodiasis (FBT)** |  |  |  |  |  |
| Thailand | Community intervention: disseminated education through posters, flipcharts, videos, group discussions (36,45,46).<br>Targeted intervention: provided training for health staff and village health volunteers (46).<br>School-based intervention: creation of school curriculum on opisthorchiasis delivered by trained teachers (46). | Individual treatment*: adults, school-aged children (36,45,46), and infected animal (46). | Community intervention: latrine installation, waste treatment. WASH practices (46). | Environment management of intermediate host (snail, cyprinid fish) elimination (45,46). | Faecal examination: human (36,45,46), and animals (45,46). |
| Lao PDR | Targeted intervention: creation of National School Health Policy (47). | MDA: school-aged children with mebendazole (47). | Targeted intervention: implemented hygiene programs in schools, emphasizing practices such as handwashing, toilet use, consuming clean food, wearing clean clothes, and maintaining a clean environment (47). | - | Faecal examination: human (47). |
| Vietnam | Targeted intervention: special training targeting nursery farmers (41,42,44).<br>Community intervention: educate household members to avoid eating raw fish and to prevent consumption of raw or dead fish by dogs, cats, and farm animals (41–43). | Individual treatment**: human in households (41–44) and animals with praziquantel (41,42,44). | Targeted intervention: sanitation program focusing on the safe disposal of human faeces and pig manure; provided hygiene instructions, including not consuming raw fish and refraining from feeding uncooked fish to animals (41–43). | Aquaculture management: pond management, snail control, and faecal pollution management (41–44). | Faecal examination: human, animals (41–44).<br>Malacology: Snail sampling to check snail density and identification (42,44). |

| Taeniasis/cysticercosis |  |  |  |  |  |
| --- | --- | --- | --- | --- | --- |
| Lao PDR | Community intervention: disseminated education through leaflets and posters such as washing hands, cooking pork, penning pigs, using latrines (33–35).<br>Community intervention: educated pig owners not to sell or consume treated pigs for at least one month after oxfendazole treatment (35). | MDA*: adult and school-aged children (triple dose albendazole) (33–35) and pigs (oxfendazole and TSOL18 vaccine) (34,35). | Community intervention: involving posters with hygiene and sanitation messages, a program on latrine usage and handwashing (33,34), and conducted latrine surveys (34). | - | Faecal examination: human (33–35) and animals (33,34). |
| Indonesia | Community intervention: disseminated education through leaflets and posters (37).<br>Targeted intervention: trained health personnel on how to diagnose and treat taeniasis (37).<br>Targeted intervention: created guidelines on the prevention and control of the disease for health workers (37). | Individual treatment of adults with praziquantel (37). | - | - | Establishment of integrated laboratory references and National Working Group (37).<br>Meat inspection: determine the distribution of infected animals (37).<br>Clinical examination: Diagnostic examination on pigs and dogs (37). |
| Philippines | Targeted intervention: capacity building for health workers to screen, interview, and perform sputum examinations on patients with a history of illness (38–40). | Individual treatment of adults with praziquantel (38–40). | - | - | Integrated surveillance and diagnostics with the National Tuberculosis Control program (38–40). |

\* This MDA targeted soil-transmitted helminthiasis, another parasitic NTD which was not focused on in this scoping review.
\*\* This treatment was conducted as part of trial study with designated group, including intervention and non-intervention groups.

### Interventions related to education

Health education is a form of activity that includes knowledge improvement and development of life skills that are beneficial to both individual and community health. Therefore, any form of education that raises awareness and targets communities or specific groups of individuals is included under this theme. Education initiatives are often conducted in conjunction with other programs such as WASH program (33,34,41–43,46,47), surveillance and diagnostics (33–39,43,45–47), ecosystem and monitoring (41–46), and treatment in humans and/or animals (33–47).

The Lawa project in Thailand emphasized education interventions in community and school settings (45,46). The authors noted that a comprehensive training program aimed at various community leaders including hospital staffs, cooks, health volunteers, village committees, and teachers proved effective in preventing Opisthorchis viverrini. The Lawa project also trained health workers to screen and treat adults, while children were targeted with a deworming program at school and a special curriculum on opisthorchiasis delivered by trained teachers. Training materials that were used in the Lawa project ranged from posters, flip charts, videos, and group discussions, which emphasized the importance of WASH practices and explain the role of animal hosts. This approach to education dissemination was also implemented in two studies to address taeniasis/cysticercosis in Indonesia (37) and Lao PDR (47). The importance of school-based interventions for health education is also highlighted by a study from Yoshida et al. (47), which found that the Ministry of Education and the Ministry of Health in Lao PDR created a National School Health Policy that included a curriculum about FBT as well as a deworming program at school.

### Interventions related to improving WASH practices

Taeniasis/cysticercosis and FBT infection in Southeast Asia are linked to cultural and community behaviours, particularly in relation to fishing, latrine use, and food preparation. Raw food consumption, such as lawar in Indonesia (37), raw pork in Lao PDR (34,35), raw freshwater crab in the Philippines (38,40), raw or fermented fish dishes in Thailand (36,45,46), and raw fish in Vietnam (41,44), present a source of infection for taeniasis and FBT infection. Therefore, WASH initiatives related to reducing the incidence of taeniasis/cysticercosis and FBT infection typically focused improving hygiene and sanitation practices, including with respect to food preparation and consumption.

WASH programs across countries in Southeast Asia were delivered alongside education and ecosystem monitoring interventions. Hygiene programs in the included studies were undertaken in Thailand (46), Lao PDR (33,34,47), and Vietnam (41–43), and aimed to educate individuals on handwashing, toilet use, and food preparation. While sanitation programs focused on conducting latrine surveys (47), installing latrines, and managing human and animal waste (41–43,46). In Lao PDR (34), the study highlighted that taeniasis/cysticercosis posters related to WASH were effective in increasing women’s knowledge about the disease, but were not successful for children and men.

### Interventions via treatment

Treatment is the administration of drugs for the purpose of promoting public health. One treatment approach for NTDs consists of mass drug administration (MDA), in which the whole population (or a particular sub-set of the population) are all targeted for treatment regardless of their infection status; another way of treatment is through medication only for infected individuals. MDA was only conducted in studies on FBT and taeniasis/cysticercosis in Lao PDR (33–35,47), while the remaining studies focused on individual treatment. In all included studies where individual treatment was administered (36–46), treatment was preceded by stool examination as part of diagnostics and surveillance in animals and/or humans.

A study in Lao PDR demonstrated the implementation of an MDA program among adults and school-aged children against taeniasis/cysticercosis. Adults were given albendazole 400 mg over three consecutive days, while all pigs received anthelmintic oxfendazole 30 mg/kg and TSOL18 vaccine. This procedure was repeated one month later (33–35). This study aimed to not only to control taeniasis/cysticercosis but also soil-transmitted helminthiasis. Although surveillance measures showed an increment of soil-transmitted helminths among humans, taeniasis prevalence in humans was reduced after the first MDA. Studies conducted in Thailand, Lawa project, also mentioned the integration of treatment for opisthorchiasis within other programs (45,46). The project joined in other control programs aimed at helminths, vector-borne, and zoonotic diseases, administering praziquantel 40 mg/kg to reservoir hosts (cats and dogs) and infected human individuals.

Studies conducted in Vietnam demonstrated the positive effects of a holistic approach to treatment, incorporating ecosystem monitoring and community education, not only for humans but also for animals, as described in more detail in the next section. Faecal examinations conducted on humans and domestic animals revealed statistically significant reductions in the density and total count of FBT eggs (41–44) after the multisectoral interventions were implemented. Despite these promising results, challenges remain including the management of free-roaming domestic animals, which can impede efforts to control the spread of disease.

### Interventions related to ecosystem monitoring

Ecosystem monitoring plays an important role in controlling parasitic zoonotic NTDs by identifying areas of high risk and informing efforts to control disease, and can also incorporate control of intermediate hosts or vectors. Aquatic environments like lakes, rivers, and local ponds are often monitored to determine the prevalence and abundance of freshwater snails, which are the intermediate hosts of numerous zoonotic trematodes, in areas where FBT is endemic. This information is used to implement preventative measures like reducing snail populations, which in turn can help curb disease.

Studies in Vietnam (41–44) looked at the strong integration of ecosystems monitoring, diagnostics, treatment, and education interventions in the domain of One Health. The ecosystem was monitored focusing on aquaculture management practices such as pond management, snail control, and faecal pollution management. Investigators trained all household members including aquaculture farmers about the basic biology of FBT. Furthermore, the farmers were given specific instructions for pond nursery management. Instructions included redirecting human and pig waste away from ponds, installing fences around ponds (to prevent domestic animals from entering pond areas), applying water filters and removing vegetation. Additionally, intervention groups and animals were examined and treated with praziquantel. Results demonstrated that the prevalence of FBT eggs among cats and dogs decreased post-intervention compared to controls. However, the result also showed that improving pond and farm management practices can be effective even without drug treatment.

### Intervention by surveillance and diagnostics

Surveillance and diagnostics are critical in mapping the necessary tools and methods for effective disease intervention, as well as providing insights into environmental factors that contribute to the infection cycle. Both surveillance and diagnostics were implemented to some degree in all included studies to evaluate intervention success and establish baseline information about taeniasis and FBT.

Faecal examination was one of the primary methods used in surveillance and diagnostic interventions. The One Health approach was specifically identified in studies where faecal examinations were conducted not only on human excrement but also on domestic animals (33,34,41–46). However, in studies in Lao PDR on FBT, diagnostics were only performed on humans (47). Studies conducted in the Philippines (38–40) emphasized the importance of routine surveillance and diagnostics for FBT integrated into health centres. A paragonimiasis control program was integrated within the National Tuberculosis Control Program due to their similarity of clinical symptoms and shared diagnostic methods. However, this integration only focused on human health and in this sense, was not an ideal example of a One Health approach. Nonetheless, the National Tuberculossis Control Program highlighted opportunities for integration and gaps for improvement. Similarly, Indonesia conducted passive surveillance at health centres and established collaboration activities with veterinary public health officials (37). This sector supported laboratory cooperation through the Bureau of Veterinary Research and Development and performed diagnostic examinations on pigs and dogs. The government also conducted meat inspections at registered slaughterhouses to determine the distribution of infected animals.

Indonesia and the Philippines faced similar challenges, such as limited personnel skills for laboratory diagnostics and program monitoring, infrastructure limitations for surveillance, and a lack of political commitment. These external factors have shown that these diseases are not prioritized in the health agenda, and no clear cross-sectoral intervention guidelines were identified.

### Challenges encountered during intervention implementation

Four studies (41,44–46) identified budgeting and monitoring as the primary challenges for implementing One Health interventions. Delivering medication treatment for humans and animals over a long period of time can be expensive, and ensuring all hosts receive treatment is difficult, especially since farm owners sometimes bring in new cats and dogs without prior notice. The extent of community coverage of MDA in humans can be substantially affected by the time of day or day of the week; the efficacy of school-based MDA can be impacted by high rates of absenteeism or low school enrolment, which can particularly negatively affect girls’ access to treatment (48). Bardosh et al. (34) also reported that the inadequate supply chain of porcine vaccines for taeniasis increased the likelihood of unsuccessful programs in Lao PDR, particularly in isolated villages. Studies also identified low personnel capacity as a hindrance to program implementation (34,37). A lack of trust in health interventions was identified as another challenge to the program’s success. In Lao PDR, some parents did not allow their children to take anthelmintic during deworming programs in school as they feared side effects, as reported by Okello et al. (35). Hygiene and sanitation practices were challenging to improve, as culturally determined behaviours continued to hinder education and WASH interventions. Bardosh et al. (34) and Ash et al. (33) found that many villagers in Lao PDR were unwilling to build or use cement latrines and continued unhygienic cooking, which increased reinfection.

Political commitment with effective coordination is critical for the sustained implementation of One Health programs. However, a study in Indonesia highlighted that government resources and priorities were not focused on taeniasis control, but other diseases for example polio, HIV/AIDS, malaria, and dengue which were supported by international agencies. Thus a more efficient way to control taeniasis was averted (37). In the Philippines, the absence of guidelines and measurements for an integrated paragonimiasis control program with the tuberculosis national program lead to low implementation rates (38). Delos Trinos et al. (40) found that MDA guidelines from the Philippines government were not aligned with recommendations from the WHO to control FBT.

## Discussion

This scoping review presents the multi/cross-sectoral engagement in controlling parasitic zoonotic NTDs and highlights the challenges of implementing One Health approaches in Southeast Asia. In addition to being implemented singly, some of the thematic interventions we identified in the review were implemented together with others, including both multisectoral and unisectoral efforts. For example, our review revealed that education interventions, specifically those emphasizing WASH practices, were frequently implemented alongside treatment interventions for disease control. However, the positive impact of even these multi-layered interventions was not always demonstrated. In Lao PDR, despite extensive implementation of health education and deworming programs for school-age children, the prevalence of Opisthorchis sp. infection remained high at 39% (47). A meta-analysis by Naqvi et al. (49) found that the impact of health education on NTDs is uncertain, although it may reduce the intensity and prevalence of schistosomiasis, another parasitic NTD, among children and adolescents. In this scoping review, health education material often consisted of posters and leaflets, but according to Hasanica et al. (50), these were ineffective tools to improve children’s knowledge about NTDs. Considering the need for behavioural change for sustainable impacts on NTDs, alternative health education tools are necessary. For example, a pilot study in Nigeria used the board game “Schisto and Ladders version 2” to increase schistosomiasis knowledge, resulting in a significant change in treatment-seeking behaviour from 0% to 65.3% (51), suggesting that gamification may be more effective than text-based media in some contexts.

It has been known the exact effect of WASH interventions on NTD prevention and control is difficult to determine due to insufficient conclusive evidence (52). A review by Garn et al. (53) found that increased latrine coverage or improvements do not necessarily result to an increase in usage. Factors influencing latrine use include latrine types, accessibility, privacy, cleanliness, hygiene amenities, functionality, and maintenance. Furthermore, the study by Bardosh et al. (34) highlights a gender-based difference in sanitary infrastructure usage due to their roles. In the study, the educational material mainly improved women’s knowledge and raised their awareness for sanitary conditions, while it barely affected men and children. Therefore, gender analysis is critically needed to explore opportunities for a more gender-responsive approach in health interventions. The WHO NTD road map also clearly highlights the importance of ensuring gender equity and human rights in all services provided for NTDs (8).

Preventive chemotherapy and treatment are key WHO-recommended interventions for NTD prevention and control, but strategies focused exclusively on humans show limited results. In Madagascar, a three-year pilot study found that mass drug administration (MDA) reduced taeniasis prevalence by 80%, however, prevalence returned to baseline levels 16 months after the final MDA round (54). This highlights the need for One Health approach that includes intermediate hosts like pigs in the intervention strategies. The included studies showed excellent results by implementing interventions on pigs by using a combination of TSOL18 vaccine, which has proven to be the most effective vaccine against T. solium, and oxfendazole to reduce source of taeniasis infection in humans. This successful approach was also reported from a study in Uganda (55) as well as using combination of TSOL16 and TSOL18 in Peru (56). Another study in Morocco suggested that EG95 vaccination on sheep reduce the level of cystic echinococcosis (57). This emphasizes the importance of employing an integrated approach rather than a siloed one to address zoonotic diseases challenges. Additionally, co-administering anthelmintic drugs to control multiple diseases also presents a promising example of integration within health sectors even though it may not be specifically multisectoral or an example of One Health.

This review also found that ecosystem monitoring was an integral part of the One Health intervention program in several studies, particularly in controlling FBT opisthorchiasis in Vietnam and Thailand (41–44,46). The studies from Vietnam demonstrated that pond modifications are an effective method of infection prevention. The results revealed that solely improving pond and farm management can decrease opisthorchiasis and clonorchiasis infection rates in humans and animals. However, a comprehensive approach is necessary, as a mathematical modelling study (58) indicates that the optimal strategy for reducing parasite infection involves both safe fish production and routine human treatment. The cost-effectiveness of ecosystem monitoring as an intervention should be considered. Implementing pond modifications and treatment demands significant commitment and resources. A farmer’s decision to adapt control measures may be influenced by socio-cultural and economic factors (59). Transitioning from traditional agricultural practices requires a fundamental shift in perspective, which can pose financial challenges for small-scale farmers. Consequently, further research into biological alternatives for controlling infections in fish is needed, along with the establishment of official guidelines on sustainable farm practices to prevent infections (60).

Continuous monitoring and evaluation of livestock, domestic animals, and humans are essential for identifying areas that need further intervention. Surveillance and diagnostics in humans and animals play a crucial role in One Health. The result of surveillance is useful to map areas or regions where interventions should be introduced, monitoring drug efficacy and resistance (61). Faecal examination, with its non-invasive nature and ease of sample collection, is frequently employed for diagnosing FBT and taeniasis infections (62,63). The study from Indonesia demonstrates successful intersectoral collaboration through passive surveillance efforts, showcasing the use of a veterinary laboratory as a reference for taeniasis detection (37). This approach highlights the importance of national financial commitment and strong institutional engagement in sustaining the One Health approach (64). The included studies in the Philippines also revealed the potential for integrating diagnostic programs for Paragonimus sp. and tuberculosis, given their shared symptoms and diagnostic methods (38–40). Although not directly related to One Health, such integration can help overcome the problem of insufficient test availability mentioned in a review by Bharadwaj et al. (65).

Integrating NTDs into a pre-existing initiative holds a potential to bolster the sustainability and efficiency of NTDs program management. Initiatives like the establishment of a regional network such as Mekong Basin Disease Surveillance has showcased significant advancements in enacting strategies for epidemic preparedness and response within a One Health framework. Although the focus of this regional network is around emerging infectious disease and epidemic threats, there lies a promising opportunity by incorporating NTDs. For instance, the case of Salmonellosis, a bacterial infection often linked to contaminated food. By focusing on hygienic practices within the food supply chain, not only can the risk of NTD transmission be minimized, but the occurrence of Salmonellosis and other foodborne NTDs can also be minimalized. Furthermore, considering the zoonotic nature of FBT, joint interventions with other diseases should be considered and tailored to each country’s disease situation, taking animal health sectors into account. For instance, Morocco implemented a combination of rabies vaccination, insecticidal collars for leishmaniasis control, and anthelmintic drugs for echinococcosis (66).This scoping review is subject to certain limitations. Our review was limited to online resources that are available in English, so there is a possibility that we have missed relevant studies that have been published in other languages or are not available electronically. Additionally, the exclusion of grey literature may have resulted in missing information from ministries, non-governmental organizations, and other sources that that share information on One Health intervention approaches through non-peer reviewed means. Despite these limitations, the discussion and findings summarized through key themes in this study certainly help to improve understanding of the One Health practices to advocate effective strategies for region-specific collaboration to tackle parasitic zoonotic NTDs.

To move forward in effectively controlling zoonotic NTDs, high-level decision-makers need to implement the One Health approach comprehensively. While there is a growing recognition of the importance of One Health in zoonotic NTD control efforts, it is crucial to define a research agenda and put it into practice through country or regional level initiatives.

Coordination of this kind has been developed in certain regions, such as ECOWAS in West Africa, and has been a call to action in the European Union.

## Conclusion

In conclusion, this scoping review highlights the importance and need of implementing One Health approaches rather than a siloed approach for controlling three parasitic zoonotic NTDs in Southeast Asia. However, the application of One Health strategies is not consistent throughout the countries in the Southeast Asia region, nor for diseases. For example, we could not find any studies on One Health strategies for echinococcosis, indicating a research gap. The review identifies five mains interventions that demonstrate the One Health domain, including education, treatment, WASH practices, ecosystem monitoring, and surveillance and diagnostics. However, there are challenges in implementing the One Health approach for controlling zoonotic NTDs, including limited resources, community engagement, coordination and collaboration, and political commitment. Therefore, substantial work remains to done. Continuous monitoring and evaluation of livestock, domestic animals, and humans are essential for identifying areas that need further intervention. Additionally, future potential strategies should consider determinants of health in NTDs while involve a multi-sectoral approach, including mainstreaming gender, equity, and human rights in NTD services. Integrating NTDs into broader initiatives like regional disease surveillance networks demonstrates potential synergies for efficient and sustainable management. Such initiatives can extend the impact of One Health practices, addressing not only NTDs but also other related health threats. This review also encourages decision-makers to establish coordination mechanisms in ASEAN level, drawing successful example like ECOWAS.

## Data Availability

All relevant data are within the manuscript and its Supporting Information files.

## Author contributions

Conceptualization: Agrin Zauyani Putri, Claire J. Standley

Data curation: Agrin Zauyani Putri, Adarsh Varghese George

Formal analysis: Agrin Zauyani Putri, Adarsh Varghese

George Investigation: Agrin Zauyani Putri, Adarsh Varghese George

Methodology: Agrin Zauyani Putri, Adarsh Varghese George, Claire J. Standley

Project administration: Agrin Zauyani Putri, Claire J. Standley, Shannon A. McMahon

Supervision: Claire J. Standley

Validation: Claire J. Standley

Visualization: Agrin Zauyani Putri

Writing – original draft: Agrin Zauyani Putri

Writing – review & editing: Agrin Zauyani Putri, Claire J. Standley, Shannon A. McMahon

## Notes

### Competing Interest Statement

The authors have declared no competing interest.

### Funding Statement

The author(s) received no specific funding for this work.

