## Supplementary material for "One Health Intervention Strategies to Address Zoonotic Neglected Tropical Diseases in Southeast Asia: A Scoping Review": S1_File

PUBMED SEARCH STRATEGY

### The Strategic Intervention for Neglected Zoonotic Diseases using One Health Approach in Southeast Asia: A Scoping Review

Eligibility criteria:

| Criterion | Inclusion | Exclusion |
|---|---|---|
| Literature focus | • Studies that include information about multisectoral/one health intervention strategies on Taeniasis/Cysticercosis, Echinococcosis, and Foodborne trematodiases (*Clonorchis spp., Opisthorchis spp., Fasciola spp., Paragonismus spp.*).<br>• Studies with a geographical focus on Southeast Asian countries: Brunei, Myanmar, Cambodia, Timor-Leste, Indonesia, Laos, Malaysia, Philippines, Singapore, Thailand, and Vietnam. | Studies that do not include information about multisectoral/one health intervention strategies. |
| Language | English | |
| Time Period | 2005 to 2020 | |
| Type of articles | Peer reviewed research papers | Articles that are abstract only and/or no retrievable full text, non-peer reviewed research papers |

**PUBMED Search Term**

| Keyword | # | MeSH/Text terms | Results |
|---|---|---|---|
| Neglected Zoonotic Diseases | 1 | Neglected Diseases[MeSH Terms] | 1,986 |
| | 2 | Tropical Medicine[MeSH Terms] | 1,541 |
| | 3 | Parasitic diseases[MeSH Terms] | 149,206 |
| | 4 | Zoonoses[MeSH Terms] | 10,986 |
| | 5 | Communicable Diseases[MeSH Terms] | 247,187 |
| Disease scopes | 6 | Taeniasis[MeSH Terms] | 2,905 |
| | 7 | Cysticercosis[MeSH Terms] | 2,435 |
| | 8 | Echinococcosis[MeSH Terms] | 6,129 |
| | 9 | "Trematode Infections"[MeSH Terms] | 12,601 |
| | 10 | Clonorchiasis [MeSH Terms] | 452 |
| | 11 | Opisthorchis [MeSH Terms] | 637 |
| | 12 | Fasciola[MeSH Terms] | 1,685 |
| | 13 | Paragonimus[MeSH Terms] | 277 |
| | 14 | (((Taeni*[Title/Abstract]) OR (cysticerc*[Title/Abstract])) OR (Teni*[Title/Abstract])) OR (neurocysticerc*[Title/Abstract]) | 7,126 |
| | 15 | ((((Echinococcos[Title/Abstract]) OR (hydatid*[Title/Abstract])) OR (hydatid cyst*[Title/Abstract])) OR (Echinococcus[Title/Abstract])) OR (Echinococcal disease[Title/Abstract]) | 8,548 |
| | 16 | ((((((((((Foodborne tremato d*[Title/Abstract]) OR (Trematodiasis[Title/Abstract])) OR (Clonorchiasis[Title/Abstract])) OR (Clonorchis[Title/Abstract])) OR (Opisthorchis[Title/Abstract])) OR (Opisthorchiasis[Title/Abstract])) OR (fascioliasis[Title/Abstract])) OR (Fasciola[Title/Abstract])) OR (liver fluke[Title/Abstract])) OR (paragonimiasis[Title/Abstract])) OR (Paragonimus[Title/Abstract]) | 4,883 |
| Southeast Asia | 17 | Asia, Southeastern[MeSH Terms] | 62,230 |
| | 18 | ((((((((((((brunei[Title/Abstract]) OR (burma[Title/Abstract])) OR | 69,250 |

| | | | |
|---|---|---|---|
| | | (myanmar[Title/Abstract])) OR (myanma*[Title/Abstract])) OR (cambodia[Title/Abstract])) OR (Timor-leste[Title/Abstract])) OR (Timor Leste[Title/Abstract])) OR (Indonesia[Title/Abstract])) OR (Laos[Title/Abstract])) OR (Malaysia[Title/Abstract])) OR (Philippines[Title/Abstract])) OR (Singapore[Title/Abstract])) OR (Thailand[Title/Abstract])) OR (Vietnam[Title/Abstract]) | |
| One Health Approach | 19 | One Health[MeSH Terms] | 451 |
| | 20 | ((((collboration[Title/Abstract]) OR (collaborat*[Title/Abstract])) OR (joint[Title/Abstract])) OR (integrat*[Title/Abstract])) OR (integration[Title/Abstract]) | 707,231 |
| Intervention | 21 | **Intervention[Title/Abstract]** | 505,188 |
| | 22 | ((((#1) OR (#2)) OR (#3)) OR (#4)) OR (#5) | 393,708 |
| Population | 23 | ((((((((((#6) OR (#7)) OR (#8)) OR (#9)) OR (#10)) OR (#11)) OR (#12)) OR (#13)) OR (#14)) OR (#15)) OR (#16) | 30,579 |
| Scope | 24 | (#17) OR (#18) | 87,732 |
| | 25 | **((#19) OR (#20)) OR (#21)** | 1,173,954 |
| All | 26 | **(((#22) AND (#23)) AND (#24)) AND (#25)** | **98** |

**Web of Science Search Term**

| Keyword | # | MeSH/Text terms | Results |
|---|---|---|---|
| Neglected Zoonotic Diseases | 1 | ((((TS=(Neglected Disease)) OR TS=(Tropical Medicine)) OR TS=(Parasitic diseases)) OR TS=(Zoonoses)) OR TS=(Communicable Diseases) | 983,419 |
| Disease scopes | 2 | (((((TS=(Taeniasis)) OR TS=(Cysticercosis)) OR TS=(Taeni)) OR TS=(*cysticerc*)) OR TS=(Teni*)) OR TS=(neurocysticerc*) | 28,843 |
| | 3 | ((((TS=(Echinococcosis)) OR TS=(Echinococcos hydatid*)) OR TS=(hydatid cyst*)) OR TS=(Echinococcus)) OR TS=(Echinococcal disease) | 11,242 |
| | 4 | ((((((((((((((((((((((TS=(Trematode Infections)) OR TS=(Clonorchiasis)) OR TS=(Opisthorchis)) OR TS=(Fasciola)) OR TS=(Paragonimus)) OR TS=(Taeni*)) OR TS=(cysticerc*)) OR TS=(Teni*)) OR TS=(neurocysticerc*)) OR TS=(Echinococcos)) OR TS=(hydatid*)) OR TS=(hydatid cyst*)) OR TS=(Echinococcus)) OR TS=(Echinococcal disease)) OR TS=(Foodborne trematod*)) OR TS=(Trematodiasis)) OR TS=(Clonorchiasis)) OR TS=(Clonorchis)) OR TS=(Opisthorchis)) OR TS=(Opisthorchiasis)) OR TS=(fascioliasis)) OR TS=(Fasciola)) OR TS=(liver fluke)) OR TS=(paragonimiasis)) OR TS=(Paragonimus) | 66,938 |
| Southeast Asia | 5 | (((((((((((((((TS=(Asia)) OR TS=(Southeastern)) OR TS=(Southeast*)) OR TS=(brunei)) OR TS=(burma)) OR TS=(myanmar)) OR TS=(myanma*)) OR TS=(cambodia)) OR TS=(Timor-leste)) OR TS=(Timor Leste)) OR TS=(Indonesia)) OR TS=(Laos)) OR TS=(Malaysia)) OR TS=(Philippines)) OR TS=(Singapore)) OR TS=(Thailand)) OR TS=(Vietnam) | 1,450,416 |
| One Health Approach | 6 | (((((TS=(One Health)) OR TS=(collaboration)) OR TS=(collaborat*)) OR TS=(joint)) OR TS=(integrat*)) OR TS=(integration) | 3,267,362 |
| Intervention | 7 | TS=(intervention) | 1,137,809 |
| Population | 8 | #2 OR #3 OR #4 | 67,788 |
| appch | 9 | #6 OR #7 | 4,201,012 |
| All | 10 | #1 AND #8 AND #9 AND #5 | 1,529 |

**Ovid Medline**

Ovid MEDLINE(R) and Epub Ahead of Print, In-Process, In-Data-Review & Other Non-Indexed Citations, Daily and Versions <1946 to May 27, 2022>

| Keyword | # | MeSH/Text terms | Results |
|---|---|---|---|
| Neglected Zoonotic Diseases | 1 | exp Neglected Diseases/ | 1972 |
| | 2 | exp Tropical Medicine/ | 1534 |
| | 3 | exp Parasitic Diseases/ | 148042 |
| | 4 | exp Zoonoses/ | 10876 |
| | 5 | exp Communicable Diseases/ | 244840 |
| Disease scopes | 6 | exp Taeniasis/ | 2889 |
| | 7 | exp Cysticercosis/ | 2421 |
| | 8 | exp Echinococcosis/ | 6076 |
| | 9 | exp Trematode Infections/ | 12504 |
| | 10 | exp Clonorchiasis/ | 448 |
| | 11 | exp Opisthorchis/ | 633 |
| | 12 | exp Fasciola/ | 1665 |
| | 13 | exp Paragonimus/ | 276 |
| | 14 | (Taeni* or cysticerc* or teni* or neurocysticerc*).mp. [mp=title, abstract, original title, name of substance word, subject heading word, floating sub-heading word, keyword heading word, organism supplementary concept word, protocol supplementary concept word, rare disease supplementary concept word, unique identifier, synonyms] | 8300 |
| | 15 | (Echinococcos or hydatid* or "hydatid cyst*" or Echinococcus or Echinococcal disease).mp. [mp=title, abstract, original title, name of substance word, subject heading word, floating sub-heading word, keyword heading word, organism supplementary concept word, protocol supplementary concept word, rare disease supplementary concept word, unique identifier, synonyms] | 7748 |

| | | | |
|---|---|---|---|
| | 16 | ("Foodborne trematod*" or Trematodiasis or Clonorchiasis or Clonorchis or Opisthorchis or Opisthorchiasis or fascioliasis or Fasciola or "liver fluke" or paragonimiasis or Paragonimus).mp. [mp=title, abstract, original title, name of substance word, subject heading word, floating sub-heading word, keyword heading word, organism supplementary concept word, protocol supplementary concept word, rare disease supplementary concept word, unique identifier, synonyms] | 4700 |
| Southeast Asia | 17 | exp Asia, Southeastern/ | 61332 |
| | 18 | brunei or burma or myanmar or myanma* or cambodia or Timor-leste or "Timor leste" or Indonesia or Laos or malaysia or philippines or Singapore or thailand or vietnam).mp. [mp=title, abstract, original title, name of substance word, subject heading word, floating sub-heading word, keyword heading word, organism supplementary concept word, protocol supplementary concept word, rare disease supplementary concept word, unique identifier, synonyms] | 78563 |
| One Health and Intervention | 19 | exp One Health/ | 430 |
| | 20 | ("One Health" or collaboration or collaborat* or joint or integrat* or integration or intervention).mp. [mp=title, abstract, original title, name of substance word, subject heading word, floating sub-heading word, keyword heading word, organism supplementary concept word, protocol supplementary concept word, rare disease supplementary concept word, unique identifier, synonyms] | 1186553 |
| Diseases | 21 | 1 or 2 or 3 or 4 or 5 | 390159 |
| | 22 | 6 or 7 or 8 or 9 or 10 or 11 or 12 or 13 or 14 or 15 or 16 | 31994 |
| Population | 23 | 17 or 18 | 85584 |
| Intervention | 24 | 19 or 20 | 1186661 |
| All | 25 | 21 AND 22 AND 23 AND 24 | **98** |
